# Can we mitigate the psychological impacts of social isolation using behavioural activation? Long-term results of the UK BASIL Urgent Public Health COVID-19 pilot randomised controlled trial and living systematic review

**DOI:** 10.1101/2022.06.20.22276641

**Authors:** Elizabeth Littlewood, Dean McMillan, Carolyn A. Chew-Graham, Della Bailey, Samantha Gascoyne, Claire Sloan, Lauren Burke, Peter Coventry, Suzanne Crosland, Caroline Fairhurst, Andrew Henry, Catherine Hewitt, Kalpita Baird, Eloise Ryde, Leanne Shearsmith, Gemma Traviss-Turner, Rebecca Woodhouse, Judith Webster, Nick Meader, Rachel Churchill, Elizabeth Eddy, Paul Heron, Nisha Hickin, Roz Shafran, Osvaldo P. Almeida, Andrew Clegg, Tom Gentry, Andrew Hill, Karina Lovell, Sarah Dexter Smith, David Ekers, Simon Gilbody

**Author notes:** Declared competing interests of authors: none.

## Abstract

**Background:** Behavioural and cognitive interventions remain a credible approach in preventing loneliness and depression. There was a need to rapidly generate and assimilate trial-based data during COVID-19.

**Objectives:** We undertook a COVID-19 parallel pilot RCT of behavioural activation for depression and loneliness [the BASIL-C19 trial ISRCTN94091479]. We also assimilate these data in a COVID-19 living systematic review [PROSPERO CRD42021298788].

**Methods:** Primary care participants (>=65 years) with long-term conditions were computer randomised to Behavioural Activation (n=47) versus care-as-usual (n=49). The single blinded primary outcome was the PHQ-9. Secondary outcomes included loneliness (De Jong Gierveld Scale). Data from the BASIL-C19 trial were included in a random effects meta-analysis of depression and loneliness.

**Findings:** The 12 months adjusted mean difference for PHQ-9 was -0.70 (95% CI -2.61 to 1.20) and for loneliness was -0.39 (95% CI -1.43 to 0.65). Secondary 12-month trial outcomes suggested evidence of benefit for behavioural activation.

The BASIL-C19 meta-analysis (13 trials) found short-term reductions in depression (standardised mean difference [SMD]=-0.31, 95%CI -0.51 to -0.11) and loneliness (SMD=-0.48, 95%CI -0.70 to -0.27). There were few long-term trials, but there was evidence of some benefit (loneliness SMD=-0.20, 95%CI -0.40 to -0.01; depression SMD=-0.20, 95%CI -0.47 to 0.07).

**Discussion:** We found a signal of effect in reducing loneliness and depression in the BASIL trial. Living meta-analysis provides strong evidence of short-term benefit for loneliness and depression.

**Clinical implications:** Scalable behavioural and cognitive approaches should be considered as population-level strategies for depression and loneliness on the basis of the living systematic review.

**Funding:** This study was funded by National Institute for Health and Care Research (NIHR) Programme Grants for Applied Research (PGfAR) RP-PG-0217-20006.

**Author summary:** *Why was this study done?:*

⍰ Older people with long-term conditions have been impacted by COVID-19 pandemic restrictions and have experienced social isolation. In turn, this puts them at risk for depression and loneliness, and these are bad for health and wellbeing. Psychosocial approaches, such as behavioural activation, could be helpful.
⍰ Trial-based evidence is needed to demonstrate if it is possible to prevent the onset, or mitigate the impact, of loneliness and depression.
⍰ There are few studies of brief psychosocial interventions to mitigate depression and loneliness, and it is important to know how emerging trial-based data adds to existing evidence.

*What did the researchers do and find?:*

⍰ There was preliminary evidence that levels of loneliness were reduced at 3 months when behavioural activation was offered.
⍰ At longer term (12-month) follow-up there were signals of ongoing positive impact.
⍰ When BASIL-C19 data were assimilated into a living systematic review there is clear evidence of impact of brief psychological interventions on depression and loneliness in the short-term. More research into the longer-term impact is needed.

*What does all this mean?:*

⍰ Behavioural activation now shows evidence of benefit which will be useful for policy makers in offering support to people who are socially isolated.
⍰ This research knowledge will be useful once the COVID-19 pandemic has passed, since loneliness is common in older populations and effective scalable solutions will be needed to tackle this problem.
⍰ As new trial-based data emerges, our living systematic review and meta-analysis will be updated since this is an area of active research.

## Introduction

The mental health of the population deteriorated during the COVID-19 pandemic ^1^. Many people reported social isolation, and the incidence of depression and anxiety particularly increased for older people and those with medical vulnerabilities ^2^. A plausible mechanism for this deterioration was that COVID-19 restrictions led to disruption of daily routines, loss of social contact and heightened isolation and increased loneliness, which are each powerful precipitants of mental ill health ^3^.

Social isolation, social disconnectedness, perceived isolation and loneliness are known to be linked to common mental health problems, such as depression in older people ^3 4^. Loneliness is a risk factor for depression and seems detrimental to physical health and life expectancy ^5^. It is recognised that strategies that, for instance, maintain social connectedness could be important in ensuring the mental health of older people ^6^, particularly during the pandemic ^3^ and in the planning for post-pandemic recovery ^7^ (including the management of people with Long Covid.

The need for research to mitigate the psychological impacts of COVID-19, particularly loneliness, was highlighted as a priority ^8^, and we responded by designing and delivering one of a small number of psychotherapy trials programmes ^9^.

Behavioural activation (BA) is an evidence-based psychological treatment that explores how physical inactivity and low mood are linked and result in a reduction of valued activity ^10^. Small scale trials of BA delivered to socially-isolated older people have produced encouraging preliminary results ^11^, but there is not yet sufficient research evidence to support whole-scale adoption, or to inform the population response to COVID-19 or in planning for post-pandemic recovery. We therefore adapted an ongoing work programme into the role of BA in multiple long term conditions in early-2020 to answer the following overarching question: **‘Can we prevent or ameliorate depression and loneliness in older people with long-term conditions during isolation?’**.

In this paper we present the long-term (12-month) results of the BASIL-C19 trial (**<u>B</u>**ehavioural **<u>A</u>**ctivation in **<u>S</u>**ocial **<u>I</u>**so**<u>l</u>**ation): a pilot randomised controlled trial (RCT) of manualised BA, adapted specifically to be delivered at scale and remotely (via the telephone or video call) for older adults who became socially isolated as a consequence of COVID-19. The long-term (12-month outcomes) complement the already-published short-term (up to 3 months) outcomes of the BASIL-C19 trial ^12^. This is a rapidly evolving area, and we therefore present the results of the BASIL-C19 trial alongside all randomised data in a prospective evidence synthesis and cumulative meta-analysis (a ‘living systematic review’) ^13^.

## Trial methods

### Study design and participants

The BASIL-C19 pilot RCT was the first and only mental health trial adopted by the National Institute for Health and Care Research (NIHR) Urgent Public Health programme (adopted on 28^th^ May 2020) ^14^. The BASIL-C19 pilot was designed to provide key information on methods of recruitment and training for intervention practitioners (hereafter BASIL Support Workers [BSWs]). The trial was registered on 9^th^ June 2020 (ISRCTN94091479) and participants were recruited between 23^rd^ June and 15^th^ October 2020. Older adults with long-term conditions were identified as being a ‘high risk group’ for loneliness and depression as a consequence of social isolation under COVID-19 restrictions. They were recruited from primary care registers in the North East of England. Eligible and consenting participants were randomised to receive either usual primary care (with signposting to resources to support mental health during COVID) from their general practice or Behavioural Activation intervention in addition to usual care. Methods, recruitment, intervention uptake, retention, experience of the BA intervention for our target population, and acceptability of the intervention are described in full in the short-term results paper ^12^.

#### Inclusion criteria

Based on the Academy of Medical Sciences definition of multimorbidity ^15^ we recruited older adults (65 years or over) with two or more physical long-term conditions (LTCs) on primary care registers in two general practices in the North East of England. Participants included those subject to English Government guidelines regarding COVID-19 self-isolation, social distancing and shielding as relevant to their health conditions and age (though this was not a requirement and these requirements changed during the study period).

#### Exclusion criteria

Older adults who had cognitive impairment, bipolar disorder /psychosis/ psychotic symptoms, alcohol or drug dependence, in the palliative phase of illness, had active suicidal ideation, were currently receiving psychological therapy, or are unable to speak or understand English.

Potentially eligible participants were telephoned and those who expressed an interest in the study were contacted by a member of the research team to determine eligibility, obtain consent and collect baseline data. Interested patients could also complete an online consent form or contact the study team directly.

### Randomisation, concealment of allocation and masking

Eligible and consenting participants were randomised 1:1 to BA intervention or usual care using simple randomisation via an automated computer data entry system, administered remotely by the York Trials Unit, University of York. Participants, general practices, study clinicians, or BSWs were not blinded to treatment allocation. Outcome assessment was by self-report, and study researchers facilitating the telephone-based outcome assessment were blind to treatment allocation.

### Intervention (Behavioural Activation)

The intervention (BA within a collaborative care framework) has been described elsewhere ^16^ and was adapted for the purposes of the BASIL-C19 trial. The main adaptation was the use of telephone delivery, and the use of functional equivalence to maintain social interactions. Intervention participants were offered up to eight sessions over a 4 to 6 week period delivered by trained BSWs, accompanied by a BASIL Behavioural Activation booklet.

Sessions were delivered by BSWs remotely via telephone or video call, according to participant preference. The first session was scheduled to last approximately one hour, with subsequent sessions lasting approximately 30 minutes.

#### Comparator (usual GP care with signposting)

Participants in the control group received usual care as provided by their current NHS and/or third sector providers. In addition, control participants were ‘signposted’ to reputable sources of self-help and information, including advice on how to keep mentally and physically well (e.g., Public Health England (PHE) ‘Guidance for the public on the mental health and wellbeing aspects of coronavirus (COVID-19)’ ^17^ and Age UK ^18^).

### Outcome measures

Demographic information obtained at baseline included: age, sex, long-term condition type, socio-economic status, ethnicity, education, marital status, and number of children.

The overarching aim of the BASIL-C19 pilot trial was to test the feasibility of the intervention and the methods of recruitment, randomisation and follow-up ^19^. The primary clinical outcome was self-reported symptoms of depression, assessed by the PHQ-9 ^20^, where higher scores initiate greater levels of depressive symptomatology. The PHQ-9 was administered at baseline, one, three and 12 months post-randomisation. Other secondary clinical outcomes measured at baseline, one, three and 12 months were health related quality of life (SF-12v2 mental component scale (MCS) and physical component scale (PCS)) ^21^, anxiety (GAD-7) ^22^, perceived social and emotional loneliness (De Jong Gierveld Scale - 11 items loneliness scale) and questions relating to COVID-19 circumstances and adherence to government guidelines ^23^. Findings from one and three month outcomes have been presented elsewhere ^12^, along with information on intervention compliance.

### Sample size & statistical analysis

#### Sample size

Sample size calculations were based on estimating attrition and standard deviation (SD) of the primary outcome. We aimed to recruit 100 participants. The intervention was delivered by BSWs and allowed for potential clustering by BSWs assuming an inter-cluster correlation (ICC) of 0.01 and mean cluster size of 15 based upon previous studies ^16^. The effective sample size was therefore 88. Anticipating 15-20% of participants would be lost to follow-up (17% in the CASPER trial of older adults ^16^), this would result in an effective sample size of at least 70 participants, which is sufficient to allow reasonably robust estimates of the SD of the primary outcome measure to inform the sample size calculation for a definitive trial ^24^.

#### Statistical analysis

This study is reported as per the Consolidated Standards of Reporting Trials (CONSORT) guideline. The flow of participants through the pilot trial is shown in a CONSORT flow diagram [Figure 1]. Differences in the clinical outcomes between the two groups were compared at 12 months. This was done using a covariance pattern, mixed-effect linear regression model incorporating all post-randomisation time points. Treatment group, time point, a treatment-by-time interaction and the baseline score of the outcome of interest were included as fixed effects, and participant as a random effect (to account for the repeated observations per participant).

**Figure 1:**
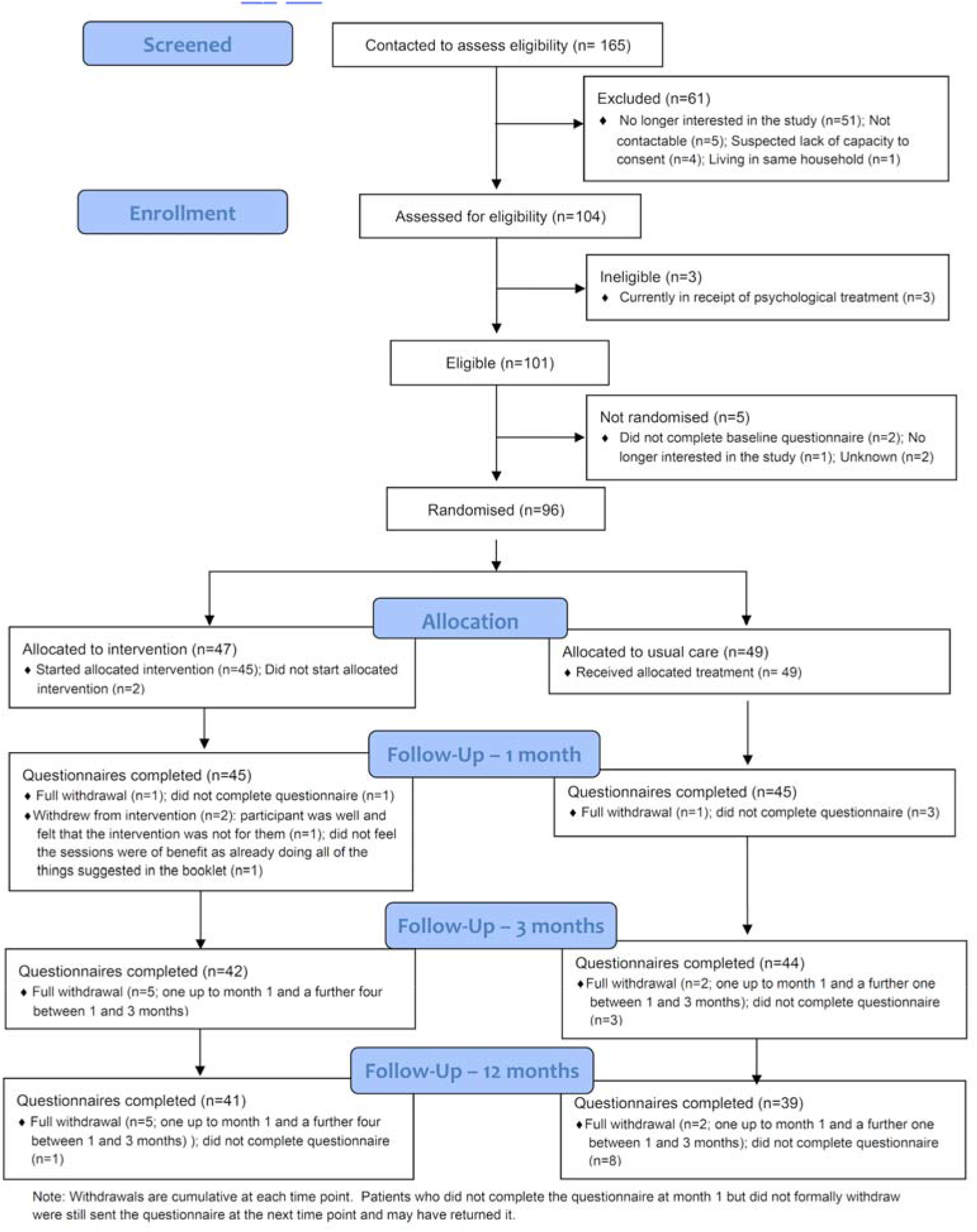
BASIL CONSORT flow diagram.

**Figures 2 & 3:**
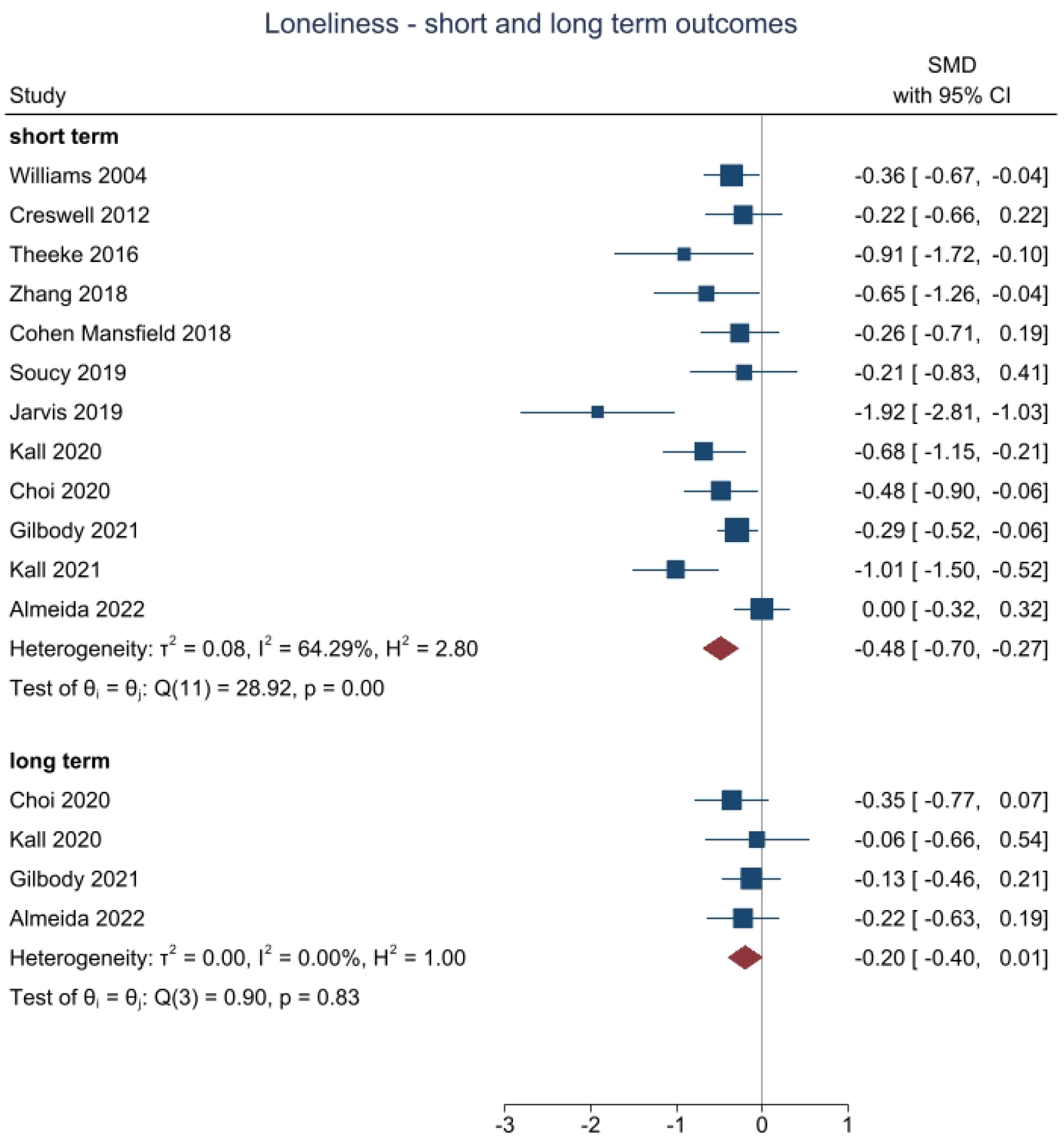
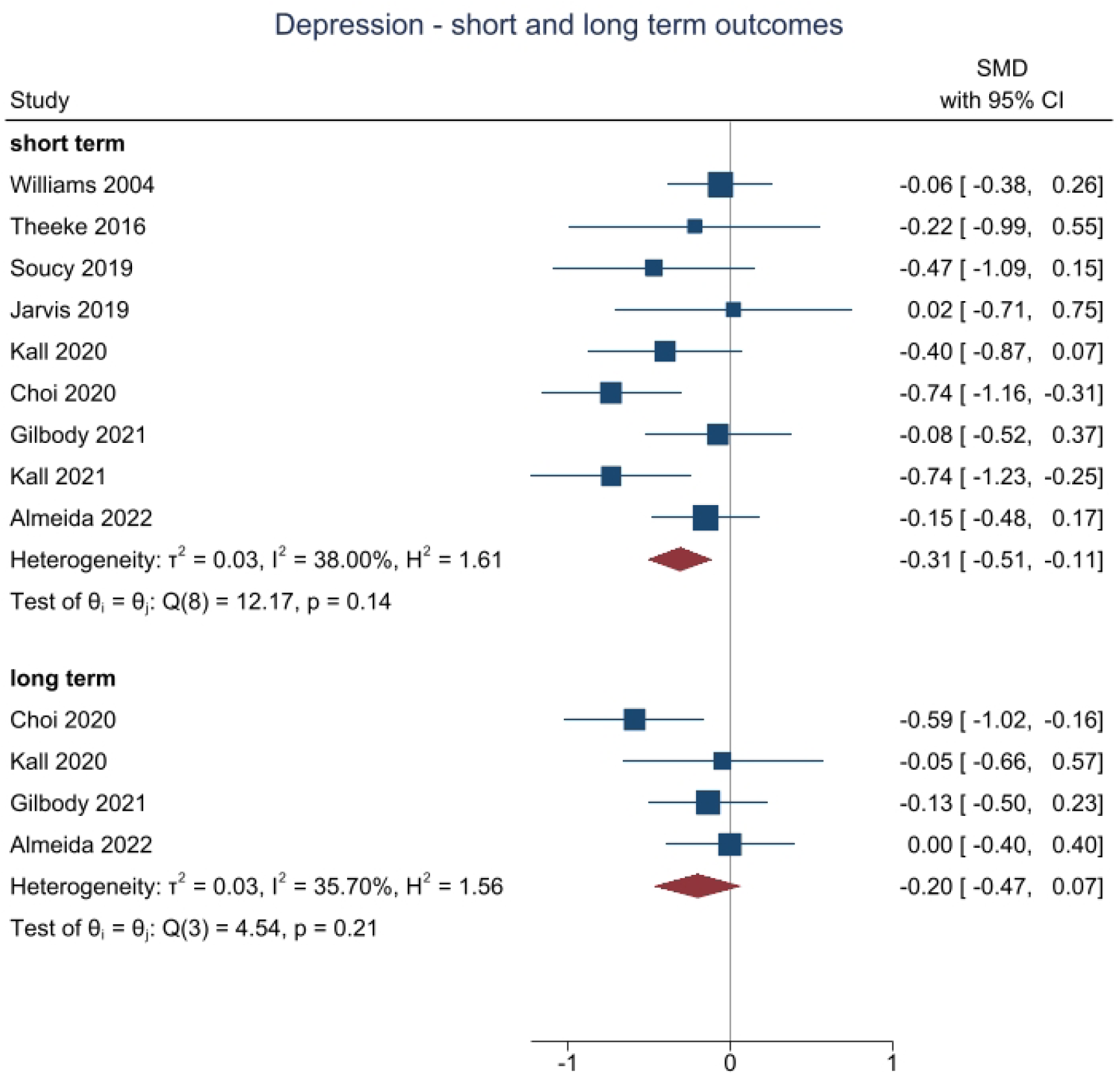
Living meta-analysis of behavioural and cognitive trials targeting loneliness and depression in socially isolated populations.

Different covariance structures were applied to the model. An unstructured covariance pattern for the correlation between the observations for a participant over time was specified in the final model based on Akaike’s Information Criterion (AIC) (smaller value preferred).

An estimate of the difference between treatment groups in all outcome measures was extracted from the models for the 12-month time point, and overall, with a 95% confidence interval (CI) as preliminary estimates of effect, but this pilot trial was not powered to show efficacy. Model assumptions were checked as follows: the normality of the standardised residuals was visually assessed using a QQ plot, and homoscedasticity by means of a scatter plot of the standardised residuals against fitted values. No concerning deviations were noted.

### Prospective meta-analysis of trial-based data

Using all trial data to February 2022 we then updated an earlier Cochrane ^25^ and non-Cochrane ^26^ meta-analysis of cognitive or behavioural interventions to prevent or mitigate loneliness and depression in adult populations in light of the BASIL-C19 results. The planned living systematic review and meta-analysis protocol was registered on the PROSPERO database (review protocol CRD42021298788).

We updated Cochrane searches of PubMED, EMBASE and PsycINFO from inception to February 2022. Eligible interventions included first, second, or third wave cognitive behavioural therapies (CBT) seeking to improve or prevent loneliness, as well as other CBT interventions where the focus is on improving common mental health problems but in which loneliness or a related construct is measured as an outcome. We studied depression and loneliness as the main outcomes of interest, under the advice of the BASIL Lived Experience Advisory Panel. We calculated a standardised mean difference (SMD) with 95% CI. SMD represents the size of the intervention effect of each study compared with the between-participant variability in outcome measurements recorded in each individual study. We categorised the post-intervention outcomes into short-term outcomes (< 6 months, including end of treatment time points), medium-term (≥6 to <12 months), and long-term outcomes (≥12 months). If a study reported follow-up outcomes at more than one time point within one of these time frames, we selected the outcome at the latest point within the time frame. We conducted a random effects meta-analysis, and included the BASIL-C19 study evidence. We tested for small study bias using Egger’s approach and test ^27^.

### Ethical approval

Ethical approval for the BASIL-C19 study was granted by Yorkshire & The Humber - Leeds West Research Ethics Committee on 23/04/2020 (The Old Chapel, Royal Standard Place, Nottingham, NG1 6FS, UK; +44 (0)207 104 8018), ref: 18/YH/0380 (approved as substantial amendment 02 under existing NIHR IRAS249030 research programme).

## Results

### Participant recruitment, characteristics and follow-up

Ninety-six participants were randomised using computer random number generation with concealment of allocation at the York Trials Unit (47 to the BA intervention group; and 49 to usual care with signposting group), of which 80 (83.3%) completed the 12-month follow-up and valid scores were available for 79 (82.3%). See Figure 1 [CONSORT flow diagram].

The mean age of randomised participants was 74 years (SD 5.5) and most were White (n=92, 95.8%). Nearly two-thirds of the sample were female (n=59, 61.5%) (Table 1), and the most common long-term health problems were cardiovascular conditions. Mean depression scores were indicative of mild depression (BA mean = 7.5, SD 6.2; usual care mean = 6.0, SD 5.6). There was reasonable balance in baseline characteristics at randomisation between the two groups.

**Table 1.**
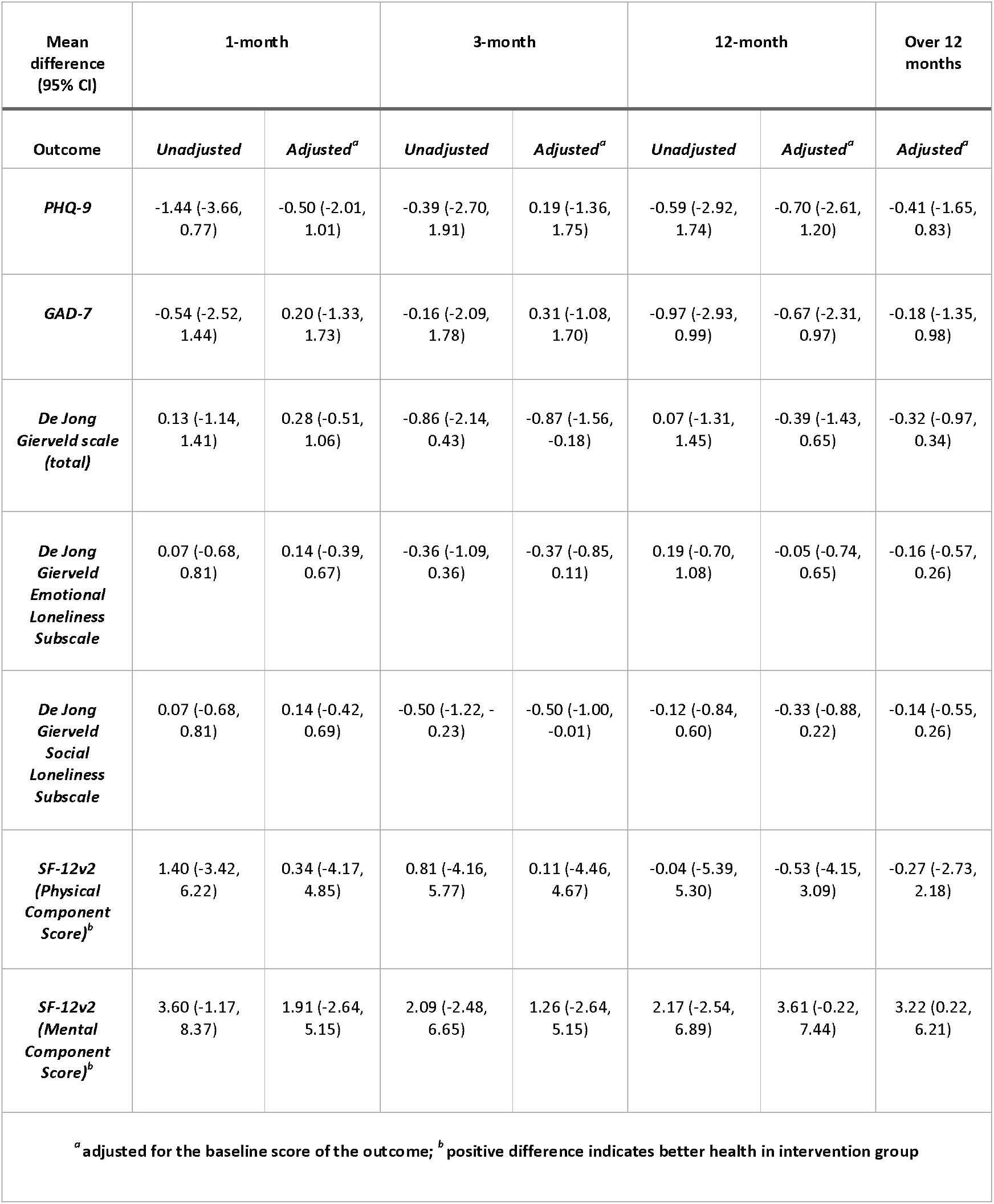
Unadjusted and adjusted mean differences between the BA and usual care groups by time point.

| Mean difference (95% CI) | 1-month |  | 3-month |  | 12-month |  | Over 12 months |
| --- | --- | --- | --- | --- | --- | --- | --- |
| Outcome | Unadjusted | Adjusted<sup>a</sup> | Unadjusted | Adjusted<sup>a</sup> | Unadjusted | Adjusted<sup>a</sup> | Adjusted<sup>a</sup> |
| **PHQ-9** | -1.44 (-3.66, 0.77) | -0.50 (-2.01, 1.01) | -0.39 (-2.70, 1.91) | 0.19 (-1.36, 1.75) | -0.59 (-2.92, 1.74) | -0.70 (-2.61, 1.20) | -0.41 (-1.65, 0.83) |
| **GAD-7** | -0.54 (-2.52, 1.44) | 0.20 (-1.33, 1.73) | -0.16 (-2.09, 1.78) | 0.31 (-1.08, 1.70) | -0.97 (-2.93, 0.99) | -0.67 (-2.31, 0.97) | -0.18 (-1.35, 0.98) |
| **De Jong Gierveld scale (total)** | 0.13 (-1.14, 1.41) | 0.28 (-0.51, 1.06) | -0.86 (-2.14, 0.43) | -0.87 (-1.56, -0.18) | 0.07 (-1.31, 1.45) | -0.39 (-1.43, 0.65) | -0.32 (-0.97, 0.34) |
| **De Jong Gierveld Emotional Loneliness Subscale** | 0.07 (-0.68, 0.81) | 0.14 (-0.39, 0.67) | -0.36 (-1.09, 0.36) | -0.37 (-0.85, 0.11) | 0.19 (-0.70, 1.08) | -0.05 (-0.74, 0.65) | -0.16 (-0.57, 0.26) |
| **De Jong Gierveld Social Loneliness Subscale** | 0.07 (-0.68, 0.81) | 0.14 (-0.42, 0.69) | -0.50 (-1.22, -0.23) | -0.50 (-1.00, -0.01) | -0.12 (-0.84, 0.60) | -0.33 (-0.88, 0.22) | -0.14 (-0.55, 0.26) |
| **SF-12v2 (Physical Component Score)<sup>b</sup>** | 1.40 (-3.42, 6.22) | 0.34 (-4.17, 4.85) | 0.81 (-4.16, 5.77) | 0.11 (-4.46, 4.67) | -0.04 (-5.39, 5.30) | -0.53 (-4.15, 3.09) | -0.27 (-2.73, 2.18) |
| **SF-12v2 (Mental Component Score)<sup>b</sup>** | 3.60 (-1.17, 8.37) | 1.91 (-2.64, 5.15) | 2.09 (-2.48, 6.65) | 1.26 (-2.64, 5.15) | 2.17 (-2.54, 6.89) | 3.61 (-0.22, 7.44) | 3.22 (0.22, 6.21) |
| <sup>a</sup> adjusted for the baseline score of the outcome; <sup>b</sup> positive difference indicates better health in intervention group |  |  |  |  |  |  |  |

### Outcome data and between-group comparisons at 12 months

Eighty randomised participants (83.3%) completed the 12-month follow-up and valid primary and secondary outcome data were available for 79 (82.3%) participants (one participant commenced the questionnaire but then felt too unwell to continue and did not complete any of the outcome measures). At 12 months, unadjusted between-group mean differences favoured the intervention for the PHQ-9, GAD-7, De Jong Social Loneliness and the SF-12 MCS, and usual care for De Jong total and the Emotional Loneliness subscale, and the SF-12 PCS. The adjusted mean difference between groups in the PHQ-9 indicated lower severity in the intervention group at 12 months (−0.70, 95% CI - 2.61 to 1.20), with an overall difference of -0.41 (95% CI -1.65 to 0.83) across all time points. The adjusted mean difference for the total De Jong Gierveld score indicated lower severity in the intervention group at 12 months (−0.39, 95% CI -1.43 to 0.65), with an overall difference of -0.32 (95% CI -0.97 to 0.34) across all time points. The direction of effect in long-term follow up was consistent, with all outcomes favouring behavioural activation, though the majority were non-significant (Table 1). For mental health-related quality of life (the SF12 mental component score) there was an overall benefit across all time points (3.22, 95% CI 0.22 to 6.21).

### Living systematic review, incorporating BASIL-C19 data with all available trials data

We identified 13 studies (including BASIL-C19) that evaluated cognitive or behavioural interventions and reported either loneliness or depression outcomes (or both) (Gilbody-BASIL 2021 ^12^, Choi ^11,28^, Pepin 2021 ^28^, Kall 2020 ^29 30^, Kall 2021 ^31^, Soucy 2019 ^32^, Williams 2004 ^33^, Zhang 2018 ^34^, Cohen-Mansfield 2018 ^35^, Cresswell 2012 ^36^, Jarvis 2019 ^37^, Theeke 2016 ^38^ and Almeida 2022 ^39^. When we pooled data, twelve studies assessed loneliness in the short-term (>=6 months) and there was strong evidence of benefit for cognitive or behavioural interventions (986 participants, SMD=-0.48, 95%CI -0.70 to -0.27, I^2^=64.3%). Four studies assessed loneliness in the long-term (>=12 months) and there was some evidence of benefit (321 participants, SMD=-0.20, 95%CI -0.40 to -0.01, I^2^ = 0%). Nine studies assessed depression in the short-term, and there was strong evidence of benefit (775 participants, SMD=-0.31, 95%CI -0.51 to -0.11, I^2^ = 38.0%). Four studies assessed depression in the long-term, at 12+ months, and although favouring cognitive or behavioural interventions the 95% CI was wider due to fewer studies reporting at this time point (324 participants, SMD=-0.20, 95%CI - 0.47 to 0.07, I^2^ = 35.7%). No studies reported medium term (>=6 to <12 month) data. In all analyses the level of between-study heterogeneity was low to moderate. Where it was possible to test for small study and publication bias, there was evidence of funnel plot asymmetry for short term loneliness (Egger test p<0.05), but not for short term depression (Egger test p= 0.76).

## Discussion

The BASIL-C19 trial is an external pilot trial, designed to test an adapted intervention and to refine trial procedures before undertaking a full-scale trial. To our knowledge, this is one of only a small number of trials undertaken during the COVID-19 pandemic to mitigate the psychological impact of the pandemic and its restrictions ^9^. We demonstrate that it was possible to trial a scalable intervention, and achieve good follow-up rates under pandemic conditions. We have previously reported the short-term outcomes ^12^, and here we present the 12-month outcomes alongside a ‘living systematic review and meta-analysis’, undertaken during the pandemic to evaluate accumulating evidence of cognitive and behavioural approaches in the prevention of depression and loneliness. Our main finding is that the BASIL-C19 pilot trial results add to a growing body of trial-based research that demonstrates that brief psychological interventions can potentially offer clinical benefit for preventing both depression and loneliness. We also demonstrate the relative absence of long-term follow up data, but note a signal of effect at 12 months and the BASIL-C19 trial is one of only three trials to assess longer term outcomes.

Research to date has shown behavioural approaches to be highly effective in the treatment of depression among older people ^10,16,40,41^ and the preliminary results of the BASIL-C19 trial support this approach under COVID-19 restrictions and in mitigating loneliness ^42^ in an at risk population.

Our pilot trial was also undertaken rapidly and during the COVID-19 pandemic in early 2020; the time elapsed between the onset of the pandemic and the recruitment of the first participant was less than 3 months. We chose to study the impact of a plausible psychosocial intervention to mitigate depression and loneliness in an at-risk population of older people with multiple long term conditions. It is important that interventions to tackle the higher rates of depression and loneliness in all age groups are also developed and evaluated.

The BASIL-C19 trial was not designed or powered to detect effectiveness, and a fully-powered pragmatic trial (BASIL+, ISRCTN63034289), is now underway to test for robust effects and replicate signals of effectiveness in important secondary outcomes such as loneliness ^43^.

The COVID-19 pandemic prompted a number of studies to understand the impacts of COVID-19,^44^ but there have been very few studies to evaluate psychosocial interventions to mitigate psychological impact ^9^. A clinical priority and policy imperative is to identify a brief and scalable intervention to prevent and mitigate loneliness, particularly in older people ^45^. The BASIL trials programme (including a living systematic review) will be informative in improving the mental health of populations in socially isolated at-risk populations after the pandemic has passed ^7^. We also emphasise that we have used, for the first time, the technique of ‘living systematic review’ to describe the impact of cognitive and/or behavioural interventions in preventing depression and loneliness in the face of social isolation and this will be updated in line with future and emerging trial based evidence. The living systematic review demonstrates that there are now multiple small-scale trials of interventions for loneliness. The strong meta-analytic signal of effect in reducing loneliness in the short term should be interpreted with some caution, since there is a potential small study bias and larger studies are needed. We note that there was a rapid rise in application of living systematic review ^13^ during the COVID pandemic, and this is one of a number of reviews that have been undertaken by the mental health research community to rapidly assimilate knowledge to inform practice and policy ^46^. An enduring legacy of the COVID pandemic might be the coupling of trials programmes with living systematic reviews, as presented in this report.

## Data Availability

The BASIL research collective is especially keen that the BASIL data contributes to prospective meta-analyses and individual patient data meta-analyses. Requests for data sharing will be considered by the independent trial steering and data monitoring committee. Full underlying (non-aggregated) data cannot be made publicly available since the ethics approval of this study does not cover openly publishing non-aggregated data.

## Contributions of the authors

SG, DE, CCG, EL, DMcM, CH, DB and SGa planned the trial, contributed to the trial design and drafted the trial protocol. EL, SG, DMcM, PC and DE led manuscript writing. EL, SG and DE oversaw the trial as chief investigators (SG, DE) and trial manager (EL), and critically revised the manuscript. SG, EL, DMcM, CCG, CH, PC, GTT, AC, TG, AHi, KL, SDS, TO and JW contributed to trial design and trial management meetings.

SG, CCG, DE, DMcM and DB designed the intervention and BSW training materials, and DB, DMcM, CCG and DE delivered the BSW training. EL led the day-to-day management of the trial, and SGa and RW were the trial coordinators. DB, SC and DMcM provided BSW clinical supervision. SGa, LB, AH, ER, LS and RW facilitated participant recruitment and follow-up data collection, and participated in trial management meetings. ER and LS delivered the BA intervention. CF, KB and CH developed the statistical analysis plan and analysed the quantitative data.

SG, DMcM, EE, PH, RS, RC & NH designed the living systematic review and are guarantors for the PROSPERO-registered review. OA provided unpublished data for the meta-analysis and is an international collaborator to the BASIL programme and the evaluation of behavioural interventions for older people.

All authors contributed to the drafts of manuscripts and read the final manuscript. The York Trials Unit act as data custodians for the BASIL-C19 trial and SG and DMcM act as data custodians for the living meta-analysis.

## Acknowledgements

We would like to thank: the participants for taking part in the trial, general practices and North East and North Cumbria Local Clinical Research Network staff for identifying and facilitating recruitment of participants, the independent Programme Steering Committee members for overseeing the study, and our BASIL Lived Experience Advisory Panel members for their insightful contributions and collaboration.

## Data sharing

A request to access these data must be made to the legal representative of the University of York. Data requestors will have to provide: i) written description and legally binding confirmation that their data use is within the scope of the study; ii) detailed written description and legally binding confirmation of their actions to be taken to protect the data (e.g. with regard to transfer, storage, back-up, destruction, misuse, and use by other parties), as legally required and to current national and international standards (data protection concept); and iii) legally binding and written confirmation and description that their use of this data is in line with all applicable national and international laws (e.g. the General Data Protection Regulation of the EU).

## References

1. Patel K, Robertson E, Kwong ASF, et al. Psychological distress before and during the COVID-19 pandemic among adults in the United Kingdom based on coordinated analyses of 11 longitudinal studies. JAMA Network open 2022; 5(4): e227629–e.

2. Steptoe A, Di Gessa G. Mental health and social interactions of older people with physical disabilities in England during the COVID-19 pandemic: a longitudinal cohort study. The Lancet Public Health 2021; 6(6): e365–e73.

3. Santini ZI, Jose PE, Cornwell EY, et al. Social disconnectedness, perceived isolation, and symptoms of depression and anxiety among older Americans (NSHAP): a longitudinal mediation analysis. The Lancet Public Health 2020; 5(1): e62–e70.

4. Lee SL, Pearce E, Ajnakina O, et al. The association between loneliness and depressive symptoms among adults aged 50 years and older: a 12-year population-based cohort study. The Lancet Psychiatry 2021; 8(1): 48–57.

5. Valtorta NK, Kanaan M, Gilbody S, Hanratty B. Loneliness, social isolation and risk of cardiovascular disease in the English Longitudinal Study of Ageing. Eur J Prev Cardiol 2018; 25(13): 1387–96.

6. Newman MG, Zainal NH. The value of maintaining social connections for mental health in older people. Lancet Public Health 2020; 5(1): e12–e3.

7. Department of Health and Social Care. COVID-19 mental health and wellbeing recovery action plan, 2021.

8. Holmes EA, O’Connor RC, Perry VH, et al. Multidisciplinary research priorities for the COVID-19 pandemic: a call for action for mental health science. Lancet Psychiatry 2020; 7(6): 547–60.

9. Gilbody S, Littlewood E, Gascoyne S, et al. Mitigating the impacts of COVID-19: where are the mental health trials? Lancet Psychiatry 2021; 8(8): 647–50.

10. Samad Z, Brealey S, Gilbody S. The effectiveness of behavioural therapy for the treatment of depression in older adults: a meta-analysis. Int J Geriatr Psychiatry 2011; 26(12): 1211–20.

11. Choi NG, Pepin R, Marti CN, Stevens CJ, Bruce ML. Improving Social Connectedness for Homebound Older Adults: Randomized Controlled Trial of Tele-Delivered Behavioral Activation Versus Tele-Delivered Friendly Visits. Am J Geriatr Psychiatry 2020; 28(7): 698–708.

12. Gilbody S, Littlewood E, McMillan D, et al. Behavioural activation to prevent depression and loneliness among socially isolated older people with long-term conditions: The BASIL COVID-19 pilot randomised controlled trial. PLoS Med 2021; 18(10): e1003779.

13. Elliott JH, Synnot A, Turner T, et al. Living systematic review: 1. Introduction—the why, what, when, and how. J Clin Epidemiol 2017; 91: 23–30.

14. NIHR. Behavioural Activation in Social Isolation (BASIL-C19). 2020. https://www.nihr.ac.uk/covid-studies/study-detail.htm?entryId=249030 (accessed 14th July 2021 2021).

15. Academy of Medical Sciences. Multimorbidity: a priority for global health research: Academy of medical sciences; 2018.

16. Gilbody S, Lewis H, Adamson J, et al. Effect of Collaborative Care vs Usual Care on Depressive Symptoms in Older Adults With Subthreshold Depression: The CASPER Randomized Clinical Trial. JAMA 2017; 317(7): 728–37.

17. Public Health England. COVID-19: guidance for the public on mental health and wellbeing. Advice and information on how to look after your mental health and wellbeing during the coronavirus (COVID-19) outbreak. 2020. https://www.gov.uk/government/publications/covid-19-guidance-for-the-public-on-mental-health-and-wellbeing (accessed 29th March 2021.

18. Age UK. Staying safe and well. 2020. https://www.ageuk.org.uk/information-advice/coronavirus/staying-safe-and-well-at-home/ (accessed 29th March 2021.

19. Eldridge SM, Chan CL, Campbell MJ, et al. CONSORT 2010 statement: extension to randomised pilot and feasibility trials. BMJ 2016; 355: i5239.

20. Kroenke K, Spitzer RL. The PHQ-9: A new depression and diagnostic severity measure. Psychiatr Ann 2002; 32: 509–21.

21. Ware JE, Kosinski M, Keller SD. SF 12: How to score the SF12 physical and mental health summary scales. Boston Mass: New England Medical Centre; 1995.

22. Spitzer RL, Kroenke K, Williams JB, Lowe B. A brief measure for assessing generalized anxiety disorder: the GAD-7. Arch Intern Med 2006; 166(10): 1092–7.

23. De Jong-Gierveld J, Kamphuls F. The development of a Rasch-type loneliness scale. Applied Psychological Measurement 1985; 9(3): 289–99.

24. Teare D, Dimairo M, Hayman A, Shephard N, Whitehead A, Walters S. Sample size requirements for pilot randomised controlled trials with binary outcomes: a simulation study. Trials 2013; 14(1): O21.

25. Eddy E, Heron PN, McMillan D, et al. Cognitive or behavioural interventions (or both) to prevent or mitigate loneliness in adolescents, adults, and older adults. Cochrane Database of Systematic Reviews (Online) 2020; [In Press].

26. Hickin N, Kall A, Shafran R, Sutcliffe S, Manzotti G, Langan D. The effectiveness of psychological interventions for loneliness: A systematic review and meta-analysis. Clin Psychol Rev 2021; 88: 102066.

27. Egger M, Davey Smith G, Schneider M, Minder C. Bias in meta-analysis detected by a simple, graphical test. BMJ 1997; 315(7109): 629–34.

28. Bruce ML, Pepin R, Marti CN, Stevens CJ, Choi NG. One Year Impact on Social Connectedness for Homebound Older Adults: Randomized Controlled Trial of Tele-delivered Behavioral Activation Versus Tele-delivered Friendly Visits. Am J Geriatr Psychiatry 2021; 29(8): 771–6.

29. Kall A, Jagholm S, Hesser H, et al. Internet-Based Cognitive Behavior Therapy for Loneliness: A Pilot Randomized Controlled Trial. Behav Ther 2020; 51(1): 54–68.

30. Käll A, Backlund U, Shafran R, Andersson G. Lonesome no more? A two-year follow-up of internet-administered cognitive behavioral therapy for loneliness. Internet interventions 2020; 19: 100301.

31. Käll A, Bäck M, Welin C, et al. Therapist-guided internet-based treatments for loneliness: a randomized controlled three-arm trial comparing cognitive behavioral therapy and interpersonal psychotherapy. Psychotherapy and Psychosomatics 2021; 90(5): 351–8.

32. Soucy I, Provencher MD, Fortier M, McFadden T. Secondary outcomes of the guided self-help behavioral activation and physical activity for depression trial. J Ment Health 2019; 28(4): 410–8.

33. Williams A, Hagerty BM, Yousha SM, Horrocks J, Hoyle KS, Liu D. Psychosocial effects of the BOOT STRAP intervention in Navy recruits. Mil Med 2004; 169(10): 814–20.

34. Zhang N, Fan Fm, Huang Sy, Rodriguez MA. Mindfulness training for loneliness among Chinese college students: A pilot randomized controlled trial. International Journal of Psychology 2018; 53(5): 373–8.

35. Cohen-Mansfield J, Hazan H, Lerman Y, Shalom V, Birkenfeld S, Cohen R. Efficacy of the I-SOCIAL intervention for loneliness in old age: Lessons from a randomized controlled trial. J Psychiatr Res 2018; 99: 69–75.

36. Creswell JD, Irwin MR, Burklund LJ, et al. Mindfulness-Based Stress Reduction training reduces loneliness and pro-inflammatory gene expression in older adults: a small randomized controlled trial. Brain Behav Immun 2012; 26(7): 1095–101.

37. Jarvis MA, Padmanabhanunni A, Chipps J. An Evaluation of a Low-Intensity Cognitive Behavioral Therapy mHealth-Supported Intervention to Reduce Loneliness in Older People. Int J Environ Res Public Health 2019; 16(7): 1305.

38. Theeke LA, Mallow JA, Moore J, McBurney A, Rellick S, VanGilder R. Effectiveness of LISTEN on loneliness, neuroimmunological stress response, psychosocial functioning, quality of life, and physical health measures of chronic illness. Int J Nurs Sci 2016; 3(3): 242–51.

39. Almeida OP, Patel H, Velasquez D, et al. Behavioural activation in nursing homes to treat depression (BAN-Dep): Results from a clustered, randomised, single-blinded, controlled clinical trial. The American Journal of Geriatric Psychiatry 2022.

40. Orgeta V, Brede J, Livingston G. Behavioural activation for depression in older people: systematic review and meta-analysis. Br J Psychiatry 2017; 211(5): 274–9.

41. Gilbody S, Brabyn S, Mitchell A, et al. Can We Prevent Depression in At-Risk Older Adults Using Self-Help? The UK SHARD Trial of Behavioral Activation. Am J Geriatr Psychiatry 2022; 30(2): 197–207.

42. Westlund E, Stuart EA. The nonuse, misuse, and proper use of pilot studies in experimental evaluation research. American Journal of Evaluation 2017; 38(2): 246–61.

43. Burke L, Littlewood E, Gascoyne S, et al. Behavioural Activation for Social IsoLation (BASIL+) trial (Behavioural activation to mitigate depression and loneliness among older people with long-term conditions): Protocol for a fully-powered pragmatic randomised controlled trial. PLoS One 2022; 17(3): e0263856.

44. Pierce M, McManus S, Jessop C, et al. Says who? The significance of sampling in mental health surveys during COVID-19. Lancet Psychiatry 2020; 7(7): 567–8.

45. O’Sullivan R, Lawlor B, Burns A, Leavey G. Will the pandemic reframe loneliness and social isolation? The Lancet Healthy Longevity 2021; 2(2): e54–e5.

46. Tasleem A, Wang Y, Li K, et al. Effects of mental health interventions among people hospitalized with COVID-19 infection: A systematic review of randomized controlled trials. General Hospital Psychiatry 2022.

